# Optimized phase-targeted auditory stimulation reduces nocturnal spikes in childhood epilepsies

**DOI:** 10.64898/2026.11.06.26367251

**Authors:** Elena C. Schmidt, Noa-Anastasia Wapp, Beatrice Alessandri, Ralf Eberhard, Debora Lederberger, Katharina Moser, Lukas Imbach, Bigna Bölsterli-Heinzle, Georgia Ramantani, Sara Fattinger, Reto Huber

## Abstract

Interictal epileptiform discharges, so called spikes, are often strongly activated during non-rapid eye movement (NREM) sleep in childhood epilepsies. In children with pronounced spike activation during NREM-sleep, spike burden can be associated with lifelong impaired cognitive functions. This spike activation is also related to alterations in sleep oscillations, specifically slow waves and spindles, which play essential roles in brain maintenance and maturation. These observations suggest that spikes may disrupt sleep-dependent brain processes that are critical for the development of cognitive functions.

Phase-targeted auditory stimulation during NREM sleep delivers brief, non-arousing tones at specific phases of ongoing slow waves to modulate sleep oscillations. Recently, our group showed that isolated, less frequent, and thus less expected, tones engage widespread thalamocortical networks, whereas repetitive, regular trains of tones promote habituation and engage more local corticocortical networks. In this study, we use these contrasting stimulation patterns for a targeted modulation of sleep oscillations and examine their interaction with epileptic spikes.

In an initial exploratory study (N = 10; age range: 7.3–16.2 years), we used a within-night block design to directly compare responses to the different stimulation patterns. Isolated tones delivered at long interstimulus intervals induced a transient increase in spike rate, whereas down-phase stimulation delivered in trains at short, regular intervals was associated with a significant reduction. We subsequently optimized the train-based paradigm and evaluated stimulation delivered throughout the night in a proof-of-concept study using a counterbalanced, between-night design (N = 5; age range: 4.4–10.7 years). Spike density decreased locally in all patients, providing convergent evidence for the effects observed in the exploratory study.

With this mechanistically guided approach, we demonstrate that repetitive down-phase auditory stimulation of slow waves results in a local reduction of epileptic spikes. This framework may provide a rational basis for targeted interventions aimed at reducing spikes and related cognitive impairments.

## Introduction

Interictal epileptiform discharges (IEDs) are often most pronounced during non-rapid eye movement (NREM) sleep. They commonly appear as spikes or sharp waves and are brief, hyper-synchronized bursts of cortical neuronal activity ^1,2^. A strong activation of IEDs during NREM sleep is a shared hallmark of wide spectrum of epilepsies where the most extreme form, the (developmental) and epileptic encephalopathies with spike-wave activation during sleep ((D)EE-SWAS) is defined by developmental regression and possible lifelong cognitive deficits ^3^. In this paediatric population, spike burden has been related to memory impairments ^4^.

Physiological NREM sleep oscillations, specifically slow waves and spindles, are linked to sleep-related memory consolidation ^5,6^ and brain development ^7^. In SWAS epilepsy patients, it has been reported that spindle density is decreased and inversely correlated to spike density ^8^ and that the precise phase-coupling of slow waves and spindles, a marker related to memory consolidation, is altered by spike density ^9^. Slow waves on the other hand have been reported to facilitate spikes, specifically in their ascending phase, both in adult focal epilepsies ^10^ and in EE-SWAS ^11^. These findings suggest that cognition, spikes and sleep oscillations are closely related; however, the causality and directionality remain unclear.

Down-phase-targeted auditory stimulation (down-PTAS) during NREM sleep allows modulating sleep oscillations noninvasively ^12^. It consists of playing short, non-arousing tones, phase-locked to the down-phase of ongoing slow waves. In healthy adults, our group recently showed that depending on the stimulation parameters, tones can influence sleep oscillations in two ways, either inducing them on the global level or suppressing them on a local level ^13^. Isolated stimuli, which are likely processed as novel sensory events, evoke a K-complex (KC), a high-amplitude, rather global, thalamocortically generated slow wave, typically followed by a sleep spindle. In contrast, repetitive stimulation delivered during the down-phase of the slow wave, induces local desynchronization of cortical neuronal activity once the brain has habituated to the stimulus, resulting in a reduction of slow-wave activity ^13,14^.

We and others have used PTAS in paediatric patients with SWAS with different stimulation paradigms ^15–17^. These studies yielded somewhat contradictory results but overall showed a potential interaction between PTAS and epileptic spikes. Based on previous literature, it therefore remains unclear which stimulation approach could be most effective in reducing epileptic spikes. One study successfully reduced epileptic spikes and proposed that isolated stimuli, by inducing K-complexes and associated sleep spindles, could reduce epileptic activity through a disruption or “hijacking” of the thalamocortical loop ^16^. Alternatively, repetitive stimulation delivered during the slow-wave down-phase (down-PTAS), which is proposed to locally desynchronize cortical activity, could counteract the hypersynchronous neuronal activity underlying epileptic spikes ^2^.

Here we aimed to differentiate between these two hypotheses and understand whether occupation of the thalamocortical loop or local cortical desynchronization leads to the reduction of epileptic spikes. To address this, we systematically assessed the effects of different interstimulus intervals (ISIs) and regularity of tones on epileptic spikes in paediatric patients with epilepsy. In an initial exploratory phase with a within-night block design, we found that repetitive down-PTAS with short ISI resulted in a local reduction of epileptic spikes. This effect was subsequently systematically tested in a two-night proof-of-concept study, in which application of whole night continuous stimulation led to a local reduction of spike activity compared to the Sham night. This confirmed that down-PTAS effectively reduces spikes locally, possibly through desynchronisation of cortical neuronal activity. Taken together, these findings demonstrate a mechanistically guided approach to effectively reduce epileptic spikes with PTAS.

## Materials and methods

### Patients and set-up

The study was approved by the local ethics committee (Kantonale Ethikkommission Zürich, KEK-ZH, BASEC 2019-02134) and was conducted in adherence to the principles outlined in the Declaration of Helsinki. Written informed consent was obtained from all patients or their guardians before their participation in the study. In this paper, we report data from two different experimental set-ups as outlined below (see also Figure 1).

**Figure 1.**
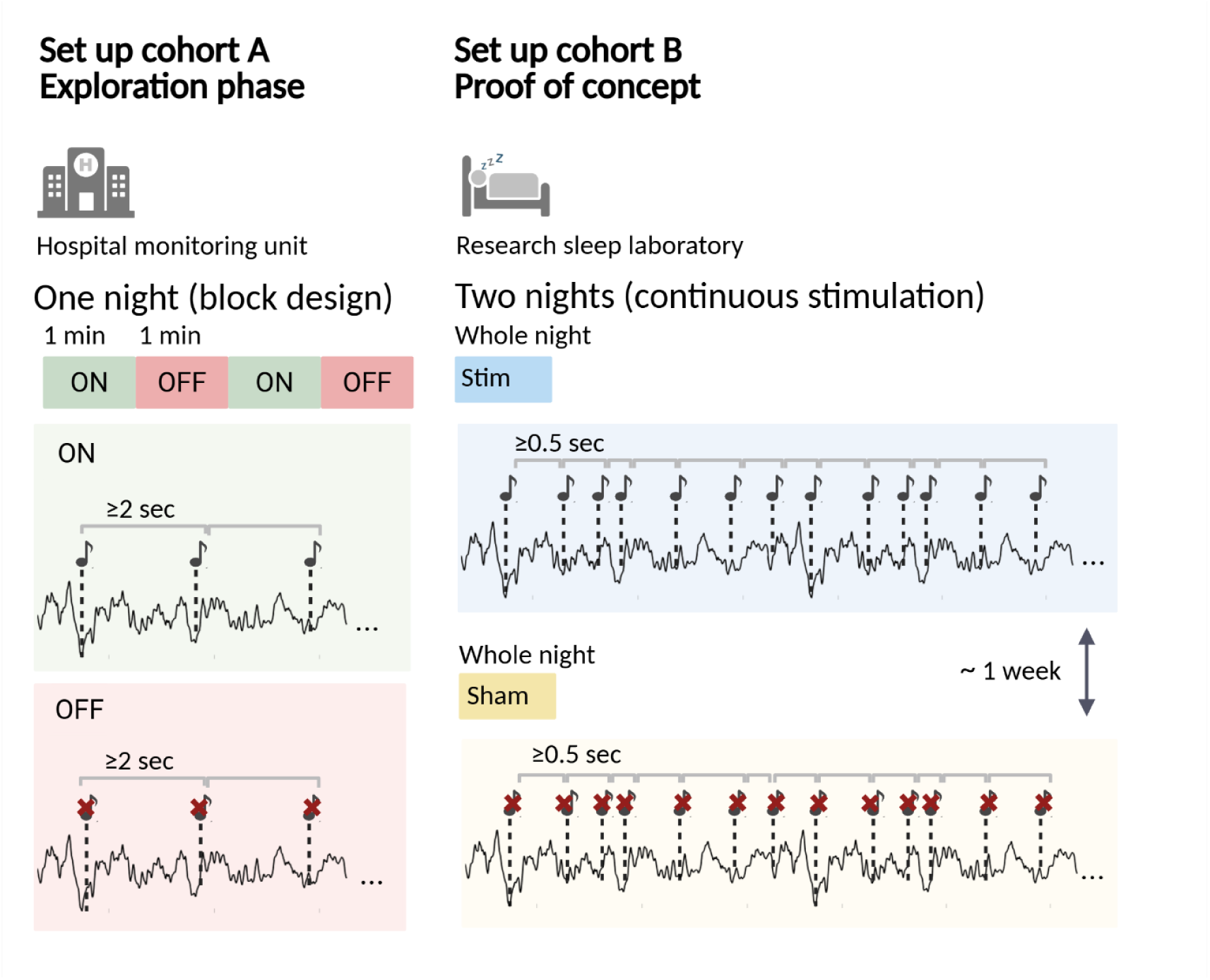
Overview of experimental set up. In cohort A, the auditory stimulation is performed during their visit in the hospital monitoring unit. Over one night the stimulation is applied in one minute on-off windows. During the ON windows, when NREM sleep is detected, stimuli are played during the down-phase of the slow wave with an interstimulus interval (ISI) of at least two seconds. Stimuli are muted in the OFF windows. In cohort B patients come to the research sleep laboratory for two nights, one true stimulation (Stim) night and one Sham (Sham) night. During the Stim night, tones are played during detected NREM sleep in the down-phase of slow waves with an ISI of at least 0.5 seconds. Stimuli are muted during the Sham night. The two nights are at least one week apart and the order is counterbalanced across patients.

For both cohorts, patients had to be between 4 and 17 years old and diagnosed with epilepsy, with the last EEG showing epileptiform discharges within 24 months prior to the recording. They were not eligible if they had an irregular sleep-wake rhythm, a hearing disorder or had travelled through 2 or more time zones in the prior month. Ongoing treatment with corticosteroids, immunosuppressants or vagus nerve stimulation, medication other than antiseizure medication and substance abuse were also exclusion criteria. Additionally for cohort B, patients with a high risk of seizures during the study night, a seizure frequency > 1/week, a seizure within 24 hours before the study night, convulsive status epilepticus since beginning of their current treatment and a history of seizure provoked by sleep deprivation were not considered eligible.

### Exploration phase - cohort A

Eligible patients were approached during their clinical visit in the epilepsy monitoring unit by the research team, in accordance with their treating physician. 18 patients were recorded in the epilepsy monitoring unit of the Children’s Hospital Zurich and the Swiss Epilepsy Centre between April 2024 and September 2025. Eight patients were excluded due to various technical problems. Ten patients were included in the final analysis (seven females, ages: 7 – 16, see Table 1).

**Table 1.** Patient characteristics. ACZ: Acetazolamide; BRV: Brivaracetam; CBD: Cannabidiol; CLB: Clobazam; DEE: developmental and epileptic encephalopathy; LCM: Lacosamide; LTG: Lamotrigine; LEV: levetiracetam; OXC: Oxcarbazepine; SeLECTS: Self-limited epilepsy with centrotemporal spikes; STM: Sulthiame; VPA: Valproic acid

| **Cohort A** |  |  |  |
|---|---|---|---|
| Patient ID | Age range | Diagnosis | Medication |
| 1A | 13 - 16 | Juvenile Absence epilepsy | LTG |
| 2A | 13 - 16 | Structural epilepsy | BRV; LCM |
| 3A | 13 - 16 | Structural epilepsy | OXC; VPA |
| 4A | 13 - 16 | Idiopathic generalized epilepsy | LTG; VPA |
| 5A | 7 - 9 | Focal epilepsy | OXC |
| 6A | 7 - 9 | DEE-SWAS | LEV |
| 7A | 7 - 9 | Generalized epilepsy | CBD; ACZ; LEV; LTG; CLB |
| 8A | 10 - 12 | Structural frontal lobe epilepsy | OXC |
| 9A | 7 - 9 | SeLECTS | None |
| 10A | 10 - 12 | Structural frontal lobe epilepsy | LCM |
| **Cohort B** |  |  |  |
| 1B | 10 - 12 | SeLECTS | STM |
| 2B | 10 - 12 | Genetic generalized epilepsy | LTG |
| 3B | 4 - 6 | SeLECTS | STM |
| 4B | 7 - 9 | SeLECTS | STM |
| 5B | 7 - 9 | SeLECTS | STM |

As part of their clinical visit, whole-night electroencephalographic (EEG) recording was performed. This was done with the Micromed Ambulatory System with 29-43 scalp electrodes, depending on clinical interest, placed according to the extended 10–20 system and sampled at 256 Hz. An additional electrode was placed close to Cz and used as reference. Electrode impedance was kept between 2 and 5 KΩ. In addition to this clinical diagnostic recording, we performed auditory stimulation during the night. The availability of 24/24-hour monitoring by trained staff made this an ideal setting to explore various stimulation parameters.

### Proof of concept - cohort B

Eligible patients were approached by their treating physician and interested patients were then contacted by the research team. Seven patients were recorded in the research sleep laboratory of the Children’s Hospital Zurich between July 2024 and May 2026. One patient dropped out due to discomfort of the net and one was excluded due to technical issues. Five patients (two females, ages: 4 – 10, see Table 1) were included in the final analysis.

Patients came to the sleep laboratory on two separate occasions for a whole-night EEG recording, one stimulation night (Stim) and one control night (Sham). To exclude potential carry-over effects, the two nights were at least seven days apart. Patients were asked to keep a regular sleep-wake cycle for three days before the recording. Adherence was checked on the evening of the recording through actimetry recordings and logs in a sleep diary. Sleep and wake time was based on their usual routine and kept the same for both recording nights. EEG was recorded using 128 channel high-density EEG (Net Amps 300 amplifier of the Geodesic EEG system (Electrical Geodesics, Inc.), Electrical Geodesics Sensor Net for long-term monitoring Hydrogel). EEG was sampled at 1000 Hz (analogue filter: 0.01–200 Hz) and referenced to the vertex (Cz). Electrooculogram (EOG) and submental electromyogram (EMG) were registered for visual sleep-stage scoring. After adjusting the net to the nasion, vertex, and mastoids, all electrodes were filled with an electrolyte gel. EEG electrode impedances were below 50 kΩ at the start of the recording and were re-checked in the morning.

### Auditory stimulation

Auditory stimulation was performed with the TOSOO Axo device, which allowed stimulation at the patient’s bedside with very minimal burden. For Cohort A, signals from an active channel, chosen to be over the epileptic focus, the respective contralateral mastoid as a reference, and the system’s ground channel as ground were fed into the Axo device with a touch-proof splitter cable. For Cohort B, an additional electrode was placed in the area of the epileptic focus and signals from both earlobe channels were fed into the Axo device, the contralateral side being used as reference and the ipsilateral as ground.

In both cohorts, signal was sampled at 250 Hz with a real-time anti-aliasing filter and a 50 Hz notch filter. The signal was immediately processed by the stimulation algorithm which recognizes NREM stages N2 and N3 in real time based on the moving average of power values (low delta, 2-4 Hz, high delta, 3-5 Hz, and high beta, 20-30 Hz) of the preceding 80s. Power thresholds were adapted for each patient based on previous thresholds used for patients with similar age and diagnosis.

A phase-locked algorithm was then applied to detect slow waves in real time and trigger stimuli between the negative zero-crossing and the trough of the ongoing slow wave. If an arousal is detected, defined as an increase in beta power within the past second, stimulation is prevented. For further information see ^18^. In the ON/Stim condition, 50 ms bursts of pink noise are being played, whereas in the control (OFF/Sham) condition, stimuli are muted.

In Cohort A, stimulations were applied in an ON-OFF window design, with a length of one minute each. The minimal inter-stimulus interval (ISI) was two seconds. Because of the clinical setting, tones were delivered over headphones with a volume of ∼50 dB.

In Cohort B, stimulation was applied continuously with a minimal ISI of 0.5 seconds. Tones were delivered with a volume of ∼50 dB through speakers attached to the bed. During one of the nights, the ‘Sham night’, tones were muted. The order of the nights was random and balanced across patients.

### Data processing

For cohort A, EEG data was exported to MATLAB, band-pass filtered between 0.5 and 40 Hz using digital filters (low-pass: passband edge 40.2 Hz, stopband 49.75 Hz, 60 dB attenuation; high-pass: passband edge 0.5 Hz, stopband 0.25 Hz, 30 dB attenuation), with passband ripple ≤0.02 and 0.05, respectively. The data was then downsampled to 128 Hz.

For cohort B, EEG data was exported to MATLAB and filtered using a zero-phase forward–backward digital filter to remove DC offset. Then it was low-pass filtered at 30 Hz using the FIRfilt plugin in EEGLAB, subsequently downsampled to 128 Hz, then high-pass filtered at 0.5 Hz.

Sleep stages were visually scored using Scoring Hero ^19^ by a trained sleep expert for consecutive 30-s epochs according to standard criteria (American Academy of Sleep medicine (AASM)) ^20^. Unclear epochs were re-evaluated with another sleep expert. After scoring, NREM sleep epochs were subjected to a semi-automatic artifact detection routine ^21^.

For all analyses, only artefact-free N2 and N3 epochs were considered. In cohort B channels with less than 90% artefact-free epochs were excluded. For topographical analyses channels below the ears were excluded and only N2 and N3 epochs which were artefact-free across all remaining channels were induced. Data was re-referenced to the linked mastoids. For one patient, signal of one mastoid had to be excluded due to bad signal quality. The data was therefore re-referenced to the linked earlobes instead.

### Spike detection

All EEG analyses were done in MATLAB R2022a. For automatic spike detection we used a previously developed method ^22^. With this method we were able to detect epileptic spikes for the whole night in all channels. In addition, to exclude potential detection biases of the algorithm, a trained technician, blinded to the condition, visually detected spikes for Cohort B in a bipolar montage in 10 second epochs. This was done for 10 min of NREM sleep in the first cycle. The segments for each patient were chosen to be comparable within the patient regarding number of stimulations and location within the night. To validate the findings of the automatic detection, the visual detection was done for a focus channel, chosen as the channel with the biggest automatically detected difference in spike density and closest to the detection electrode. A spike–wave index (SWI) was then calculated by dividing the number of seconds affected by spike waves by the number of seconds analysed, multiplied by 100 ^23^.

### Spindle detection

We used an automatic spindle detector, developed by Kramer and colleagues ^8^ to detect spindles in EEG signal with epileptic spikes. Briefly, the spindle detector estimates a latent state (the probability of a spindle) using three features: the Fano factor (estimated for data FIR filtered between 3 and 25 Hz), normalized power in the spindle band (9–15 Hz) and normalized power in the theta band (4–8 Hz). The spindle detector estimates the probability of a spindle in 0.5 s intervals (0.4 s overlap). To be considered a spindle, the probability had to be over 0.95 and the duration of at least 0.5 s. Spindles that were less than 1 s apart were merged. A subset of the detected spindles was confirmed visually for each patient. Only spindles in artefact-free, NREM 2 and 3 epochs were considered for analysis. The spindle detector is available at https://github.com/Mark-Kramer/Spindle-Detector-Method.

### Slow wave detection

Slow waves were detected using an existing algorithm ^24^. Artefact-free NREM 2 and 3 sleep data was bandpass filtered (3 dB cut-offs: 0.5 Hz and 4 Hz; Chebyshev Type II IIR filter; filter order: 2, using filtfilt) and all peaks, troughs, and zero-crossings were identified. Slow waves were defined as the time between two negative zero-crossings, with a duration of 0.5 to 2 s. As we were mostly interested in high-amplitude KC-like waves, we only analysed the top 25% of slow waves of peak-to-trough amplitude in each channel and excluded the top 5% to account for potential artefacts.

### EEG power calculation

For power calculations in cohort B, EEG data were re-referenced to the average signal across all retained channels. Power spectral density (PSD) was computed for each epoch using Welch’s method, with 4-s Hanning windows, 50% overlap, and a frequency resolution of 0.25 Hz. For slow-wave activity, mean PSD was calculated across the 0.5–4 Hz frequency range, whereas spindle activity was quantified as mean PSD across the 9–14 Hz frequency range. These epoch-wise values were then averaged across all included epochs, resulting in one mean band-specific PSD value per channel for each participant and condition.

### Estimation of phase precision and definition of region of interest

We estimated the phase precision of the stimuli, like previously done by our group ^14,25^. Briefly, the pre-processed EEG data was filtered in the slow wave range (low-pass filtered at 4 Hz). Phase values corresponding to the times of stimulation within artifact-free NREM (N2+N3) epochs were then extracted by applying the Hilbert function to the EEG data of each channel. The extracted phase values were then clustered into one of twelve 30° bins, between the two negative zero crossings. The percentage of values falling within each specific phase bin was then calculated. Stimuli were considered in the down-phase of the slow wave when they occurred in bin 2-4, equalling the period before and around the slow wave trough.

We defined a region of interest (ROI) in cohort A for statistical comparisons across patients with different locations of the detection electrode. This was done to ensure that for each patient we analysed a region with enough stimuli in the down-phase of the slow wave and enough epileptic spikes. The ROI was therefore defined as the overlap of the channels with at least 50% of stimuli in the down phase of the slow wave and at least four spikes per minute. An example of this can be found in Supplementary Figure 1.

### Statistical analysis

Statistical analyses for cohort A were done in RStudio 2026.01.2. To investigate the effect of stimuli on large-amplitude slow waves, spindles and spikes, we modelled their event probability after a stimulus (ON) compared to a muted stimulus (OFF). Generalized linear mixed-effects models (binomial logistic regression) were used to model binary event occurrence on single trial data as a function of condition (on or off stimulus), timepoint, and the interaction effects of the predictors. Participant was included as a random intercept with a random slope for condition. Because event rate was low, counts were binarized to indicate the presence versus absence of at least one event per observation window. ‘ON’ condition and time period 1 (T1; 500 ms before the stimulus until the stimulus) were used as baseline. T2-6 refer to the time between stimulus onset and 2.5 seconds after the stimulus in 500 ms steps. Post hoc estimated marginal means comparisons were computed to quantify condition differences at each timepoint. All reported p-values of the post-hoc contrast tests are Holm-corrected for multiple comparisons.

Additionally, we performed a generalized linear mixed-effects model (binomial logistic regression) to predict binary spike occurrence based on slow wave occurrence, condition, and their interaction. Participant was included as a random intercept with a random slope for condition.

Due to the lower sample size in cohort B, for paired comparisons between the Stim and Sham conditions, difference scores were calculated for each participant (Stim − Sham). Effect size was quantified using Cohen’s d, calculated as the mean paired difference divided by the standard deviation of the paired differences.

## Results

### Exploration phase - Cohort A

Recent work from our group showed that depending on regularity of the tones and the length of the ISI, auditory stimuli elicit different responses ^13,14^. More precisely, it was proposed that isolated (long ISI), therefore novel and surprising, stimuli are more likely to induce a KC followed by a spindle. Repetitive down-phase stimulation with shorter ISI, when the brain has habituated to the stimulus, induces a reduction of local slow wave activity, which might reflect a reduction of cortical neuronal synchronization. Based on these results we chose long stimulation windows (1 minute) and a medium ISI (at least 2 seconds) to ensure both types of stimulation in this cohort. Stimuli that had an ISI of at least 5 seconds to the previous stimulus were considered isolated stimuli. Stimuli with an ISI smaller than 3.5 seconds were considered part of a train. For analysis, the first five stimuli of a train were not considered, as the effect of the train stimulation emerges after the fifth stimulus ^14^. We then post-hoc divided the stimuli of Cohort A into isolated stimuli (mean number of stimuli ± SD; ON 195 ± 114.9, OFF 209.9 ± 117.3) and trains of stimuli (ON 392.7 ± 343.4, OFF 393.9 ± 400.6), similarly to ^13,14^ (see Supplementary Figure 2).

The comparison of spike density between ON and OFF windows showed no significant difference, therefore only the responses to the different types of stimuli are shown.

### Stimulus-locked response of isolated stimuli

To investigate the stimulation effect of isolated stimuli on large-amplitude slow waves, spindles and spikes, we modelled their probability after a stimulus (ON), compared to a muted stimulus (OFF). The effect on spikes is based on counts the predefined ROI (see methods and supplementary figure 1), for spindles and slow waves it was based on averaged counts from channels Fz and Cz. The full outcome of the models can be found in the supplementary table 1-6.

As shown in Figure 2 (left part) isolated stimuli increase the probability of high amplitude slow waves. This effect was most pronounced within the first 1.5 seconds as indicated by significant greater probability of large slow waves in the ON versus OFF condition (all *P* < .01) in the post-hoc comparisons (Table S1, S2).

**Figure 2.**
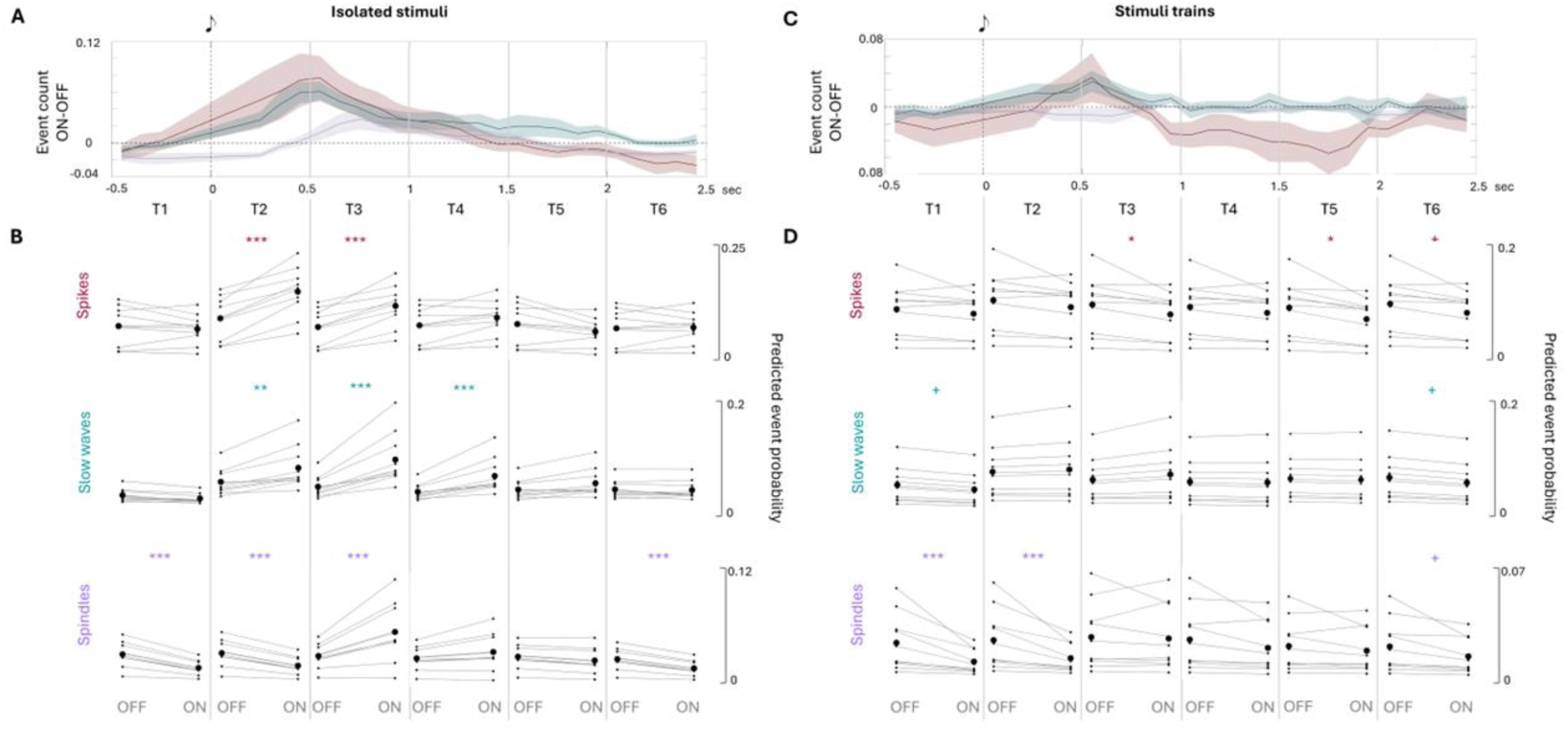
Spikes, spindles, and slow waves over time after isolated vs. trains of stimuli. Spikes, spindles, and slow waves were automatically detected across all channels using established algorithms (see methods for more details). Figure 2a *and 2c* show the time course of the events. Event counts were computed in overlapping 10 ms windows (2 ms step) spanning −0.5 to +2.5 s relative to stimulus onset for each channel and trial. The solid lines (red = spikes; blue = slow waves, purple = spindles) show the participant-averaged time course difference between ON and OFF stimuli. The shaded areas indicate the standard error of the mean. Spikes are shown in the predefined ROI. Spindles and slow waves are averaged across Fz and Cz channels for isolated stimuli, as it was previously shown that this area shows the biggest stimulation effect ^14^, and in the ROI for trains of stimuli, as we expected a local effect. Figure 2b and d show the predicted probabilities from generalized linear mixed-effects models (binomial logistic regression) for each outcome and time point. Predictions are shown separately for the ON and OFF conditions for each patient, with paired observations connected by lines. Significance stars refer to the outcome of post hoc estimated marginal means comparisons (Holm-corrected for multiple comparisons * *p* < 0.05, ** *p* < 0.01, and *** *p* < 0.001 (+ *p* < 0.1, for significance at trend level).

For spindle occurrence, post hoc comparisons showed a significantly greater spindle probability in the ON than the OFF condition at 1 second after isolated stimuli (*P* < .001), whereas spindle probability was lower in the ON condition during the first 500 ms and at 2 seconds after stimulation (all *P* < .001) (Figure 2B, and Table S3, S4).

Spike probability was increased in the ON compared to the OFF condition during the first second after an isolated stimulus (P < .001) (Figure 2B, Table S5, S6).

Since most of our patients show spike-wave complexes in their EEG we cannot fully exclude that the simultaneous stimulus-locked increase of both spikes and slow waves in cohort A is not driven by pathological spike-wave complexes. However, when excluding all slow waves that are preceded by a spike (Bölsterli Heinzle et al., 2014), to exclude spike-wave complexes, we observe the same pattern of evoked slow waves, but in reduced numbers (Supplementary Fig. 3).

As expected from previous literature and consistent with an auditory evoked KC response, the model showed an increase in the slow wave rate immediately after an isolated stimulus, followed by an increased spindle rate. In parallel, isolated stimuli were associated with a transient increase, rather than a reduction, in epileptic spike probability (Figure 2A, B).

### Spike likelihood is modulated by slow wave presence

Since we observed a simultaneous increase of spike and slow wave rate time-locked to the stimulus (Figure 2A), we investigated whether the presence of an evoked slow wave modulates the probability of a spike. Previous literature interprets the stimulus-locked high-amplitude slow wave as an evoked KC, equivalent to the auditory-evoked N550 component ^13^. We therefore considered big KC-like slow waves to be evoked when they happened 200-800 ms after an isolated stimulus. We then modelled spike probability in the same time period based on slow wave occurrence and stimulation condition. The presence of a slow wave was associated with a higher probability of a spike after a stimulus (OR = 1.23, 95% CI: 1.07, 1.43, *P* < .01). A similar association was observed in the OFF condition, indicating that both evoked and spontaneous slow waves were associated with increased spike probability (Table S13,14).

### Spikes preferentially occur before the slow wave trough

Intracranial recordings have shown that spikes preferentially occur in the down going slope before the slow-wave trough, coinciding with maximal neuronal synchrony ^10,11^. To explore this in our data, we divided the evoked slow waves into eight bins. Consistent with these findings, our exploratory analysis showed that the majority of spikes (62.5%) occurred during the descending slope of slow waves (Supplementary Figure 4).

### Stimulation in trains is associated with a reduction in spike probability

We then examined stimulus-locked responses after a train of stimuli. Slow waves, spindles and spikes were counted within the ROI (see methods and Supplementary Figure 1), as it was shown that the effects of repetitive auditory stimulation are most pronounced close to the detection electrode ^14,26^. The full outcome of the models can be found in Supplementary table *7-12*.

For slow wave occurrence, post hoc comparisons indicated no significant difference at any timepoint. Slow-wave probability was lower in the ON condition before the stimulus and at 2 seconds after the stimulus at trend level (*P* = .056) (Figure 2, right side and table S7, S8).

### Isolated stimuli and trains of stimuli elicit different responses

We observed a reduction of spindle likelihood before the stimulus for both stimulus types in cohort A. This is not visible if we only look at the first stimulus in each ON window, meaning that there was no other stimulus for at least one minute. We therefore interpret the reduction before the stimulus as the spindle refractory period ^27^ induced by spindles evoked by the previous stimulus.

Overall, these results point towards two distinct mechanisms of how isolated stimuli and stimuli in trains interact with the spikes (Figure 2). Isolated stimuli are likely to induce big KC-like slow waves that have been shown to increase spike-likelihood ^10,11^. Down-PTAS applied in trains on the other hand, reduce spikes locally, potentially through local cortical desynchronisation ^26^.

#### Proof of concept – Cohort B

Presumably due to the mixed effects of isolated stimuli and trains of stimuli, we did not see any effects when comparing spike rate between ON and OFF windows in cohort A. Additionally, the analysis of isolated and train stimuli separately revealed that train stimuli reduce spike rate significantly. In cohort B, we therefore used an optimized train stimulation protocol with the goal to reduce spike rate consistently.

In this cohort, the auditory stimulation was applied continuously time-locked to the slow wave down-phase every time a slow wave was detected, meaning without ON/OFF windows and a shorter minimal ISI (0.5 seconds) allowing for longer trains of stimuli and less isolated stimuli (mean number of stimuli ± SD; all stimuli Sham: 3715.6 ± 1761.5, single stimuli Sham: 339.4 ± 206.6, all stimuli stim: 4253.6 ± 1447.6, single stimuli stim: 423.8 ± 264.9).

### Spikes are reduced across the whole night

*Figure 3* shows the topographical distribution of the spike density for each patient in the stimulation and the Sham night. When comparing the two nights for all five patients the difference shows a clear local reduction of the spike rate in the stimulation night, with the biggest effect visible close to where the detection electrode was located. Averaged across all channels the spike density (spikes/min) in the Sham night was 5.34 ± 2.66 and 4.62 ± 2.30 in the Stim night (average ± SD), resulting in a 14.1% (Cohen’s d = 0.83) decrease of spike density in the Stim night. To estimate the local reduction, we averaged the spike density in five channels surrounding the detection electrode. This revealed a spike density of 10.39 ± 1.88 in the Sham night and 5.72 ± 2.39 in the stim night, equalling a reduction of 43.84% (Cohen’s d = 3.18) in the stim night. These results show that stimulation in trains across a whole night achieves a global spike reduction that is most pronounced in the area of the detection electrode.

**Figure 3.**
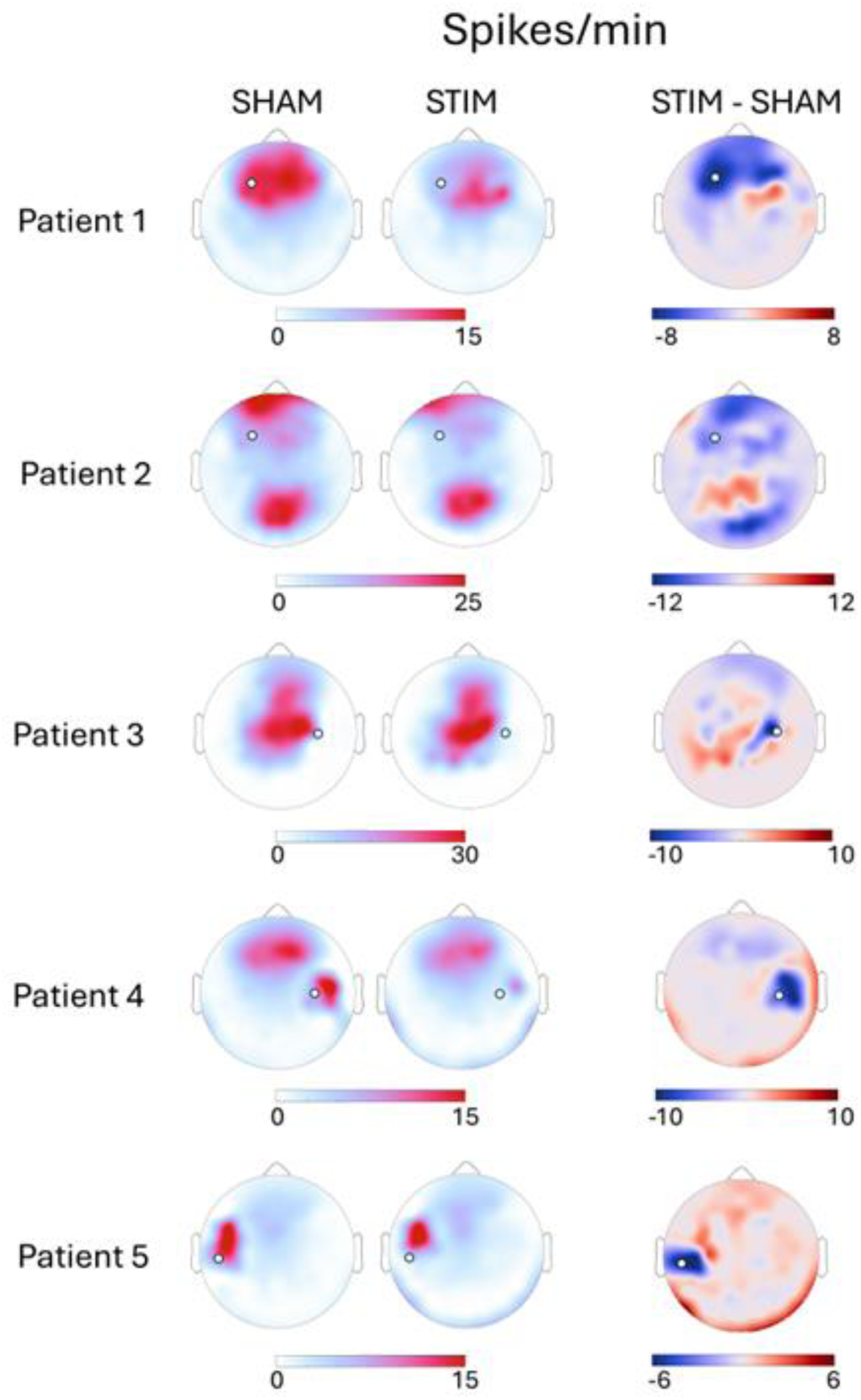
Topographical comparisons between Stim and Sham night. Spikes were automatically detected with an established algorithm ^22^ in each channel and spike density was calculated as number of events/sleep minute. The left column shows the topographical distribution in the ‘Sham’ night where no tones were played and the middle column shows the ‘STIM’ night where tones were played. The right column shows the absolute difference between the two nights (Stim - Sham). The white circle indicates the location of the detection electrode, where the slow-wave phase was detected.

### Manual spike count confirms the results

To put the results into clinical context, spikes were blindly marked by an expert manually. We then the spike wave index (SWI) in the respective focus channel (Supplementary figure 5). All five patients showed a reduction during the Stim night and spike rate was reduced during the Stim night compared with the Sham night overall (Sham: 53.60 ± 14.86%; STIM: 47.80 ± 14.45%, Cohen’s d = 1.36). These results confirm that the reduction is also detectable using the clinically established manual spike count. Importantly, it also shows that the reduction cannot be attributed to a detection bias of the automatic spike detection.

### Slow wave and spindle power is reduced in the detection channel

We then analysed power changes in the slow wave and in the spindle range, to investigate potential mechanisms for the observed spike reduction. On an individual level we did not see any clear topographical patterns between the two nights. To quantify local slow wave and spindle power changes, we then averaged the power in the focus channel for each night. As it was previously shown that power can be dominated by the response to isolated stimuli ^14^, we only considered epochs without any isolated stimuli. The results reveal a reduction of power for the slow wave reduction (Cohen’s d = 1.18) and for the spindle reduction (Cohen’s d = 1.39) in the detection channel (Supplementary figure 6).

## Discussion

The exploratory phase of the project shows that auditory stimuli evoke different responses depending on their regularity and ISI. Specifically, isolated stimuli evoked the typically observed pattern of a global, high-amplitude KC-like slow wave followed by a spindle. This response was accompanied by a transient increase in spikes predominantly around the slow wave through (Fig 2. A-B, Supplementary figure 4). Repetitive regular stimulation targeting the down phase of slow waves on the other hand, decreases spike probability and, at trend level, spindle and slow wave probability after the stimulus (Fig 2. C-D). Applying these findings to the next phase, as a proof of concept we applied stimulation optimized for repetitive regular stimulation with short ISI for a whole night and compared the spike rate to a Sham night with muted stimuli. In all five patients we saw a clear reduction of spikes in their epileptic focus, most pronounced in the area where the slow wave phase was detected (chosen to be over the epileptic focus) (Fig 3). This method therefore offers the possibility for a non-invasive way to reduce epileptic spikes during sleep.

In the recently described dual-process model of auditory stimulation ^13^, it was proposed that isolated, therefore novel, stimuli have a high likelihood of evoking a KC followed by a spindle. In this context, the globally distributed N550 component of the KC has been interpreted as the electrophysiological signature of a highly synchronized cortical off-state resulting from a transient reduction on tonic thalamocortical excitation caused by sustained inhibitory activity of thalamic reticular nucleus neurons ^28^. One previous study successfully reduced epileptic spikes with isolated stimuli and hypothesized that inducing K-complexes and associated sleep spindles, could reduce epileptic activity through a disruption or “hijacking” of the thalamocortical loop ^16^. In our results however we see an increase in spike density after isolated stimuli. We therefore hypothesize that in patients with pronounced IEDs during NREM sleep, the highly synchronous neuronal states evoked by isolated stimuli might facilitate spike generation. In accordance with this hypothesis it has previously been shown in intracranial data across different epilepsy populations, in children and adults, that (high amplitude) slow waves increase spike likelihood ^10^ and that spikes couple to the ongoing slow wave in its down-phase ^11^. In our data we show that the presence of a high-amplitude slow wave, both evoked and spontaneous, significantly increases the likelihood of a spike. Moreover, consistent with previous studies, we show in an exploratory analysis that spikes occur most frequently during the transition from the up-phase to the down-phase of the slow wave, a period of maximal cortical synchrony during which neuronal populations collectively enter the cortical off-state ^10^. Overall, our data strongly suggests that the typical evoked response from an isolated stimulus increases spike likelihood, likely through increased thalamocortical synchronisation.

In our data, in both cohorts, we observed that repetitive stimuli reduce epileptic spike likelihood significantly. In cohort A we observed a transient decrease after a stimulus, whereas in cohort B we observed this effect over a whole night. It has been proposed for decades that epileptic spikes can result from over-synchronous sleep oscillations ^2,4^. We therefore hypothesize that our stimulation reduces the synchrony of slow waves, and by that reduce the spike rate. However, the topographical effect of the repetitive stimulation on slow waves (data not shown) is not as clear as the effect on spikes. According to the dual-process model, repetitive down-phase stimulation, once the brain has habituated to the stimulus, results in local reduction of slow waves, interpreted as local cortical desynchronisation. The locality of the effect may be explained by the spatial variability of slow-wave activity across the scalp, restricting the observable reduction to the cortical area where the tone is delivered during the slow-wave down-phase ^13^. With the main goal to reduce the spikes, we chose the detection electrode based on the epileptic focus. Depending on the location, in some patients the local effects will thus not be in the area of maximal slow wave activity. This could mean that the magnitude of the induced effects is too small to detect reliably on the single subject level. In this data we therefore cannot conclude if the observed spike reduction is exclusively mediated by a reduction of slow wave activity, previously described in healthy populations ^26^, or if there is an underlying mechanism directly interacting with the spikes. To answer this question definitively, recordings with simultaneous thalamic and cortical electrodes over the epileptic focus would be needed to directly observe the effect of the stimulation on spikes in the location of their generation.

In cohort A, isolated stimuli were followed by an increase in spindle likelihood, consistent with previous findings showing a similar temporal relationship between stimulation and spindle occurrence (e.g. Leach et al., 2024). In contrast, stimulation in trains in both cohorts indicated a possible reduction in both spindle likelihood and spindle power. The interpretation of how the modulation of spindles is linked to the modulation of spikes is complicated by their bidirectional relationship reported in previous studies. Over the course of a whole night, spike and spindle density have been shown to be negatively correlated (e.g., Kramer et al., 2021; Schmidt et al., 2026), and Wodeyar and colleagues ^11^ demonstrated that cortical IEDs propagating to the thalamus can suppress spindle generation over the following seconds, providing a potential mechanistic explanation for this relationship. Conversely, spikes have also been shown to evoke spindle activity ^16^, and sigma power has been reported to positively correlate with spike activity ^29–31^. Thus, whether the changes in spindle activity observed here contribute to, or result from, the modulation of spikes remains unclear. Importantly, our stimulation effects cannot be considered independently of slow waves, as stimuli were delivered during the slow-wave down-phase and both stimulation paradigms interacted with slow-wave activity. Disentangling the contribution of spindles from that of slow waves will therefore require approaches that allow spindle activity to be modulated more selectively, such as spindle-targeted auditory stimulation ^32^ or temporal interference stimulation ^33,34^. Such approaches could help determine whether spindles play a causal role in the observed modulation of spike activity.

Both cohorts encompassed a range of diagnoses, including both focal and generalized epilepsies. If the mechanisms underlying spike generation and propagation differ between aetiologies, this heterogeneity complicates the interpretation of how stimulation interacts with epileptic activity. In particular, spikes may arise and propagate through cortical as well as thalamocortical networks ^11,35,36^. If the stimulation in trains primarily disrupts cortical synchrony, one might therefore expect a stronger effect on cortically than on thalamocortically generated spikes. We attempted to separate cohort A based on aetiology, the interpretation however was severely limited by the small sample size. In our data we therefore cannot determine if the effects of stimulation extend beyond spike generation and also influence the propagation of epileptic activity. Disentangling these effects is further complicated by the fact that, based on scalp EEG, we cannot determine whether the slow waves during which stimulation was delivered are primarily cortical or thalamocortical in origin ^37^. Thus, the observed effects could reflect modulation of cortical activity, thalamocortical interactions, or both. Larger samples allowing subgroup analyses based on diagnosis and aetiology, together with intracranial recordings, will be necessary to distinguish between these possibilities and to test the underlying mechanisms.

Our results come with certain limitations. While the location of the detection electrode in the epileptic focus area allows us to personalize the stimulation to each patient, it also makes group effects difficult to quantify and interpret. In addition, the sample size might be too small to detect subtle effects on a group level. As mentioned above, different aetiologies and scalp EEG does not allow for any mechanistic explanation of the observed stimulation effect. Finally, cognitive and clinical outcomes are missing to investigate whether the spike reduction has beneficiary effects on cognition and the overall epilepsy condition.

Nevertheless, in this study we show that epileptic spikes can be effectively reduced with continuous down-phase auditory stimulation during sleep. In the exploratory phase of the study this effect was only visible in the time immediately after the stimulus. Applying this protocol to the whole night we see the reduction in all five patients. Overall, PTAS shows great potential as a non-invasive tool to reduce the burden of epileptic spikes in children. While our approach is mechanistically guided, we cannot exclude that different stimulation parameters would be equally effective. Therefore, more studies are needed to carefully assess the optimal stimulation parameters and the effect of the stimulation over a longer period of time. Especially in more severe forms of SWAS, such as (developmental) epileptic encephalopathy with SWAS ((D)EE-SWAS), future studies would need to assess PTAS in comparison to currently available treatment options and its effect on cognitive functions.

## Data availability statement

The data is not publicly available because it contains information that could compromise the privacy of the patients. Access to the data, however, will be possible following a data sharing agreement combined with ethics approval on an individual user basis. The authors are happy to help researchers to access to this dataset.

## Acknowledgements

Figure 1 was created in BioRender. Schmidt, E. (2026) https://BioRender.com/1l86xna. This work was funded by the Swiss National Science Foundation (Project 179443) and the Swiss League Against Epilepsy. We thank Margarete Hertzsch for her valuable input to the figures and all patients and their families for participating in the study.

