## Supplementary Material for "Optimized phase-targeted auditory stimulation reduces nocturnal spikes in childhood epilepsies"

**Supplementary result tables**

**Table S1**

Slow waves after isolated stimuli

| **Predictor** | **Estimate (β)** | **OR** | **95% CI OR** | **p-value** |
| --- | --- | --- | --- | --- |
| (Intercept) | -4.45 | 0.01 | [0.01, 0.02] | **<.001** |
| Off Condition | 0.37 | 1.45 | [0.97, 2.18] | 0.072 |
| Timepoint 2 | 1.66 | 5.25 | [3.86, 7.14] | **<.001** |
| Timepoint 3 | 1.89 | 6.64 | [4.90, 8.99] | **<.001** |
| Timepoint 4 | 1.41 | 4.09 | [2.99, 5.60] | **<.001** |
| Timepoint 5 | 1.15 | 3.17 | [2.29, 4.37] | **<.001** |
| Timepoint 6 | 0.70 | 2.01 | [1.43, 2.84] | **<.001** |
| Off: Timepoint 2 | -0.87 | 0.42 | [0.28, 0.63] | **<.001** |
| Off: Timepoint 3 | -1.32 | 0.27 | [0.18, 0.41] | **<.001** |
| Off: Timepoint 4 | -1.17 | 0.31 | [0.20, 0.48] | **<.001** |
| Off: Timepoint 5 | -0.69 | 0.50 | [0.33, 0.77] | **<.01** |
| Off: Timepoint 6 | -0.32 | 0.73 | [0.47, 1.14] | 0.167 |

**Table S2**

Post-hoc tests slow waves after isolated stimuli

| **Contrast** | **Estimate (β)** | **OR** | **95% CI OR** | **p-value (Holm corrected)** |
| --- | --- | --- | --- | --- |
| Timepoint 1 On-Off | -0.37 | 0.69 | [0.46, 1.03] | .144 |
| Timepoint 2 On-Off | 0.49 | 1.64 | [1.24, 2.16] | **<.01** |
| Timepoint 3 On-Off | 0.94 | 2.57 | [1.94, 3.41] | **<.001** |
| Timepoint 4 On-Off | 0.80 | 2.22 | [1.62, 3.03] | **<.001** |
| Timepoint 5 On-Off | 0.32 | 1.38 | [1.01, 1.88] | .126 |
| Timepoint 6 On-Off | -0.05 | 0.95 | [0.68, 1.32] | .745 |

**Table S3**

Spindles after isolated stimuli

| **Predictor** | **Estimate (β)** | **OR** | **95% CI OR** | **p-value** |
| --- | --- | --- | --- | --- |
| (Intercept) | -4.63 | 0.01 | [0.00, 0.02] | **< .001** |
| Off Condition | 0.94 | 2.57 | [1.75, 3.78] | **<.001** |
| Timepoint 2 | 0.20 | 1.22 | [0.84, 1.76] | 0.301 |
| Timepoint 3 | 1.49 | 4.46 | [3.28, 6.06] | **<.001** |
| Timepoint 4 | 0.97 | 2.63 | [1.90, 3.64] | **<.001** |
| Timepoint 5 | 0.55 | 1.73 | [1.23, 2.45] | **<.01** |
| Timepoint 6 | 0.02 | 1.02 | [0.69, 1.50] | .922 |
| Off: Timepoint 2 | -0.14 | 0.87 | [0.56, 1.36] | .543 |
| Off: Timepoint 3 | -1.53 | 0.22 | [0.15, 0.32] | **<.001** |
| Off: Timepoint 4 | -1.05 | 0.35 | [0.23, 0.53] | **<.001** |
| Off: Timepoint 5 | -0.62 | 0.54 | [0.35, 0.83] | **<.01** |
| Off: Timepoint 6 | -0.21 | 0.81 | [0.51, 1.29] | .378 |

**Table S4**

Post-hoc tests spindles after isolated stimuli

| **Contrast** | **Estimate (β)** | **OR** | **95% CI OR** | **p-value (Holm corrected)** |
| --- | --- | --- | --- | --- |
| Timepoint 1 On-Off | -0.94 | 0.39 | [0.26, 0.57] | **<.001** |
| Timepoint 2 On-Off | -0.80 | 0.45 | [0.31, 0.65] | **<.001** |
| Timepoint 3 On-Off | 0.58 | 1.79 | [1.32, 2.44] | **<.001** |
| Timepoint 4 On-Off | 0.11 | 1.12 | [0.81, 1.55] | .509 |
| Timepoint 5 On-Off | -0.32 | 0.72 | [0.51, 1.02] | .132 |
| Timepoint 6 On-Off | -0.73 | 0.48 | [0.32, 0.71] | **<.001** |

**Table S5**

| **Predictor** | **Estimate (β)** | **OR** | **95% CI OR** | **p-value** |
| --- | --- | --- | --- | --- |
| (Intercept) | -2.43 | 0.08 | [0.07, 0.11] | **< .001** |
| Off Condition | 0.04 | 1.04 | [0.86, 1.26] | .701 |
| Timepoint 2 | 0.66 | 1.94 | [1.73, 2.17] | **<.001** |
| Timepoint 3 | 0.43 | 1.53 | [1.36, 1.72] | **<.001** |
| Timepoint 4 | 0.20 | 1.21 | [1.08, 1.37] | **<.01** |
| Timepoint 5 | -0.09 | 0.92 | [0.81, 1.04] | .172 |
| Timepoint 6 | 0.05 | 1.05 | [0.93, 1.19] | .429 |
| Off: Timepoint 2 | -0.53 | 0.59 | [0.50, 0.69] | **<.001** |
| Off: Timepoint 3 | -0.50 | 0.61 | [0.53, 0.72] | **<.001** |
| Off: Timepoint 4 | -0.24 | 0.79 | [0.67, 0.93] | **<.01** |
| Off: Timepoint 5 | 0.12 | 1.12 | [0.94, 1.33] | .191 |
| Off: Timepoint 6 | -0.09 | 0.92 | [0.77, 1.09] | .324 |

Spikes after isolated stimuli

**Table S6**

| **Contrast** | **Estimate (β)** | **OR** | **95% CI OR** | **p-value (Holm corrected)** |
| --- | --- | --- | --- | --- |
| Timepoint 1 On-Off | -0.04 | 0.96 | [0.79, 1.17] | 1.00 |
| Timepoint 2 On-Off | 0.49 | 1.63 | [1.36, 1.96] | **<.001** |
| Timepoint 3 On-Off | 0.46 | 1.58 | [1.31, 1.91] | **<.001** |
| Timepoint 4 On-Off | 0.20 | 1.22 | [1.01, 1.48] | .154 |
| Timepoint 5 On-Off | -0.15 | 0.86 | [0.71, 1.04] | .370 |
| Timepoint 6 On-Off | 0.05 | 1.05 | [0.86, 1.27] | 1.00 |

Post-hoc tests spikes after isolated stimuli

**Table S7**

Slow waves after trains of stimuli

| **Predictor** | **Estimate (β)** | **OR** | **95% CI OR** | **p-value** |
| --- | --- | --- | --- | --- |
| (Intercept) | -3.55 | 0.29 | [0.02, 0.05] | **<.001** |
| Off Condition | 0.25 | 1.28 | [1.06, 1.55] | **<.01** |
| Timepoint 2 | 0.73 | 2.07 | [1.76, 2.45] | **<.001** |
| Timepoint 3 | 0.59 | 1.81 | [1.53, 2.15] | **<.001** |
| Timepoint 4 | 0.38 | 1.46 | [1.22, 1.74] | **<.001** |
| Timepoint 5 | 0.48 | 1.61 | [1.35, 1.91] | **<.001** |
| Timepoint 6 | 0.27 | 1.31 | [1.10, 1.57] | **<.01** |
| Off: Timepoint 2 | -0.28 | 0.76 | [0.60, 0.95] | **.019** |
| Off: Timepoint 3 | -0.38 | 0.68 | [0.54, 0.87] | **<.01** |
| Off: Timepoint 4 | -0.19 | 0.83 | [0.65, 1.06] | .137 |
| Off: Timepoint 5 | -0.17 | 0.84 | [0.66, 1.07] | .159 |
| Off: Timepoint 6 | -0.06 | 0.97 | [0.76, 1.25] | .837 |

**Table S8**

Post-hoc tests slow waves after trains of stimuli

| **Contrast** | **Estimate (β)** | **OR** | **95% CI OR** | **p-value (Holm corrected)** |
| --- | --- | --- | --- | --- |
| Timepoint 1 On-Off | -0.25 | 0.78 | [0.65, 0.94] | 0.056 |
| Timepoint 2 On-Off | 0.03 | 1.03 | [0.89, 1.19] | 1.000 |
| Timepoint 3 On-Off | 0.14 | 1.15 | [0.98, 1.34] | 0.372 |
| Timepoint 4 On-Off | -0.06 | 0.94 | [0.80, 1.11] | 1.000 |
| Timepoint 5 On-Off | -0.08 | 0.93 | [0.79, 1.09] | 1.000 |
| Timepoint 6 On-Off | -0.22 | 0.80 | [0.68, 0.95] | 0.056 |

**Table S9**

Spindles after trains of stimuli

| **Predictor** | **Estimate (β)** | **OR** | **95% CI OR** | **p-value** |
| --- | --- | --- | --- | --- |
| (Intercept) | -5.34 | 0.00 | [0.00, 0.01] | **<.001** |
| Off Condition | 0.97 | 2.63 | [1.77, 3.90] | **<.001** |
| Timepoint 2 | 0.19 | 1.21 | [0.86, 1.70] | .267 |
| Timepoint 3 | 1.05 | 2.85 | [2.13, 3.81] | **<.001** |
| Timepoint 4 | 0.76 | 2.13 | [1.58, 2.89] | **<.001** |
| Timepoint 5 | 0.63 | 1.88 | [1.38, 2.57] | **<.001** |
| Timepoint 6 | 0.40 | 1.49 | [1.08, 2.06] | **.016** |
| Off: Timepoint 2 | -0.13 | 0.88 | [0.58, 1.33] | .542 |
| Off: Timepoint 3 | -0.88 | 0.42 | [0.28, 0.60] | **<.001** |
| Off: Timepoint 4 | -0.64 | 0.53 | [0.36, 0.77] | **<.001** |
| Off: Timepoint 5 | -0.74 | 0.47 | [0.32, 0.70] | **<.001** |
| Off: Timepoint 6 | -0.50 | 0.61 | [0.41, 0.91] | **.016** |

**Table S10**

Post-hoc tests spindles after trains of stimuli

| **Contrast** | **Estimate (β)** | **OR** | **95% CI OR** | **p-value (Holm corrected)** |
| --- | --- | --- | --- | --- |
| Timepoint 1 On-Off | -0.97 | 0.38 | [0.26, 0.56] | **<.001** |
| Timepoint 2 On-Off | -0.84 | 0.43 | [0.30, 0.63] | **<.001** |
| Timepoint 3 On-Off | -0.09 | 0.91 | [0.65, 1.27] | .600 |
| Timepoint 4 On-Off | -0.33 | 0.72 | [0.51, 1.02] | .195 |
| Timepoint 5 On-Off | -0.23 | 0.80 | [0.56, 1.14] | .433 |
| Timepoint 6 On-Off | -0.47 | 0.62 | [0.43, 0.91] | .052 |

**Table S11**

| **Predictor** | **Estimate (β)** | **OR** | **95% CI OR** | **p-value** |
| --- | --- | --- | --- | --- |
| (Intercept) | -2.46 | 0.09 | [0.06, 0.13] | **<.001** |
| Off Condition | 0.10 | 1.10 | [0.95, 1.29] | .202 |
| Timepoint 2 | 0.15 | 1.17 | [1.07, 1.27] | **<.001** |
| Timepoint 3 | 0.03 | 1.03 | [0.94, 1.13] | .515 |
| Timepoint 4 | 0.05 | 1.05 | [0.96, 1.15] | .308 |
| Timepoint 5 | -0.02 | 0.98 | [0.89, 1.07] | .605 |
| Timepoint 6 | 0.02 | 1.02 | [0.93, 1.11] | .709 |
| Off: Timepoint 2 | 0.03 | 1.03 | [0.91, 1.17] | .619 |
| Off: Timepoint 3 | 0.10 | 1.10 | [0.97, 1.25] | .121 |
| Off: Timepoint 4 | 0.02 | 1.01 | [0.90, 1.16] | .719 |
| Off: Timepoint 5 | 0.14 | 1.15 | [1.02, 1.31] | **.028** |
| Off: Timepoint 6 | 0.09 | 1.09 | [0.96, 1.24] | .173 |

Spikes after trains of stimuli

**Table S12**

Post-hoc tests spikes after trains of stimuli

| **Contrast** | **Estimate (β)** | **OR** | **95% CI OR** | **p-value (Holm corrected)** |
| --- | --- | --- | --- | --- |
| Timepoint 1 On-Off | -0.10 | 0.91 | [0.78, 1.05] | 0.259 |
| Timepoint 2 On-Off | -0.13 | 0.88 | [0.75, 1.02] | 0.259 |
| Timepoint 3 On-Off | -0.20 | 0.82 | [0.70, 0.95] | **0.049** |
| Timepoint 4 On-Off | -0.12 | 0.88 | [0.76, 1.03] | 0.259 |
| Timepoint 5 On-Off | -0.24 | 0.79 | [0.67, 0.91] | **0.011** |
| Timepoint 6 On-Off | -0.19 | 0.83 | [0.71, 0.96] | 0.062 |

**Table S13**

Spike probability based on slow wave occurrence after isolated stimuli

| **Predictor** | **Estimate (β)** | **OR** | **95% CI OR** | **p-value** |
| --- | --- | --- | --- | --- |
| (Intercept) | -1.75 | 0.17 | [0.11, 0.27] | **<.001** |
| Slow wave | 0.21 | 1.23 | [1.07, 1.43] | **<.01** |
| Off condition | -0.55 | 0.58 | [0.38, 0.87] | **<.01** |
| Slow wave: off condition | 0.28 | 1.32 | [1.05, 1.65] | **.018** |

| **Contrast** | **Estimate (β)** | **OR** | **95% CI OR** | **p-value** |
| --- | --- | --- | --- | --- |
| On Condition  No slow wave –slow wave | -0.21 | 0.81 | [0.70, 0.94] | **<.01** |
| Off Condition  No slow wave –slow wave | -0.49 | 0.62 | [0.52, 0.73] | **<.001** |

**Table S14**

Post-hoc tests spike probability based on slow wave occurrence after isolated stimuli

**Table S15**

Slow waves after isolated stimuli (without ‘spike waves’)

| **Predictor** | **Estimate (β)** | **OR** | **95% CI OR** | **p-value** |
| --- | --- | --- | --- | --- |
| (Intercept) | -4.34 | 0.01 | [0.01, 0.02] | **<.001** |
| Off Condition | 0.23 | 1.25 | [0.87, 1.81] | 0.232 |
| Timepoint 2 | 1.12 | 3.06 | [2.24, 4.19] | **<.001** |
| Timepoint 3 | 1.24 | 3.47 | [2.55, 4.72] | **<.001** |
| Timepoint 4 | 0.93 | 2.53 | [1.83, 3.48] | **<.001** |
| Timepoint 5 | 0.87 | 2.39 | [1.73, 3.30] | **<.001** |
| Timepoint 6 | 0.41 | 1.51 | [1.07, 2.14] | **.020** |
| Off: Timepoint 2 | -0.50 | 0.61 | [0.40, 0.93] | **.021** |
| Off: Timepoint 3 | -0.89 | 0.41 | [0.27, 0.63] | **<.001** |
| Off: Timepoint 4 | -0.89 | 0.41 | [0.26, 0.65] | **<.001** |
| Off: Timepoint 5 | -0.69 | 0.50 | [0.32, 0.79] | **<.01** |
| Off: Timepoint 6 | -0.26 | 0.77 | [0.48, 1.24] | 0.282 |

**Table S16**

| **Contrast** | **Estimate (β)** | **OR** | **95% CI OR** | **p-value (Holm corrected)** |
| --- | --- | --- | --- | --- |
| Timepoint 1 On-Off | -0.23 | 0.80 | [0.55, 1.15] | .232 |
| Timepoint 2 On-Off | 0.28 | 1.32 | [1.02, 1.70] | **.034** |
| Timepoint 3 On-Off | 0.67 | 1.95 | [1.49, 2.54] | **<.001** |
| Timepoint 4 On-Off | 0.66 | 1.93 | [1.43, 2.62] | **<.001** |
| Timepoint 5 On-Off | 0.47 | 1.59 | [1.19, 2.14] | **<.01** |
| Timepoint 6 On-Off | 0.03 | 1.03 | [0.75, 1.43] | .842 |

Post-hoc tests slow waves after isolated stimuli (without ‘spike waves’)

**Supplementary Figures**

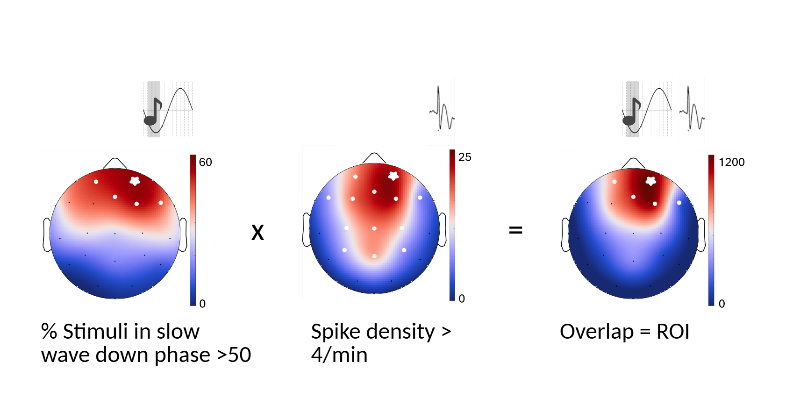

***Supplementary Figure 1* – Data preparation in cohort A**

Since the detection electrode differs between patients according to their epileptic focus, a region of interest is determined to perform group analyses. This procedure is shown for one example patient. The left part shows the percentage of stimuli in the slow wave down phase for each channel (for details see the method section and the supplementary material). All channels reaching at least 50% are selected and marked with a white dot. The middle panel shows the spike density for each channel. Here, all channels with at least four spikes per minute (on average) are selected and marked in white. The intersection of selected channels fulfilling both criteria (marked with white circles) defines the selected region of interest (ROI) for this patient. For visualisation purposes, the right panel shows the multiplication of the two other values for each channel. The detection electrode is marked with a white star in all panels.

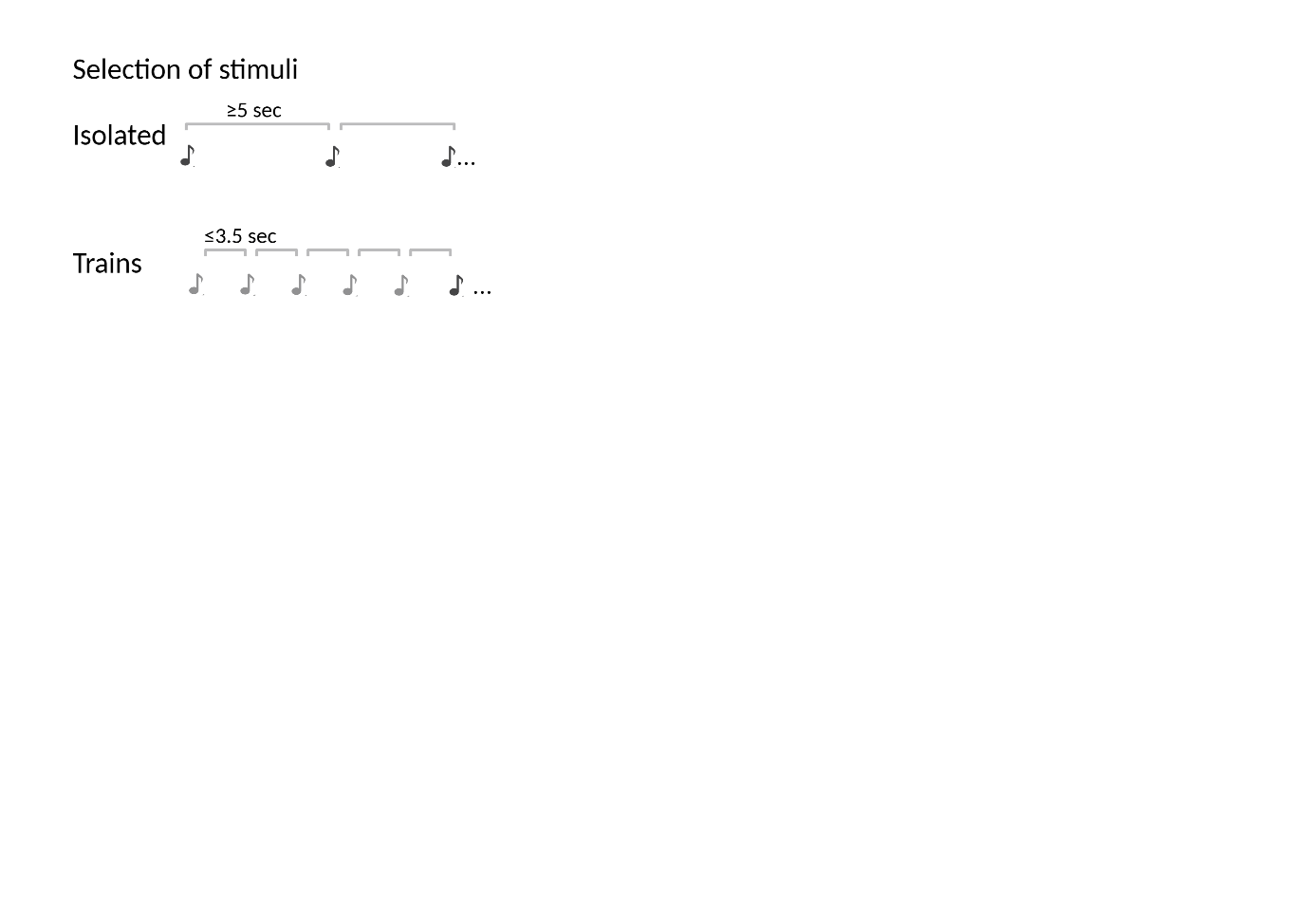

***Supplementary Figure 2* – Selection of stimuli in cohort A**

Post-hoc, stimuli in cohort A were sorted into isolated stimuli and trains of stimuli. Stimuli that had an ISI of at least 5 seconds to the previous stimulus were considered isolated stimuli. Stimuli with an ISI smaller than 3.5 seconds were considered part of a train. For analysis, the first five stimuli of a train were not considered, based on the results by Leach et al., 2025.

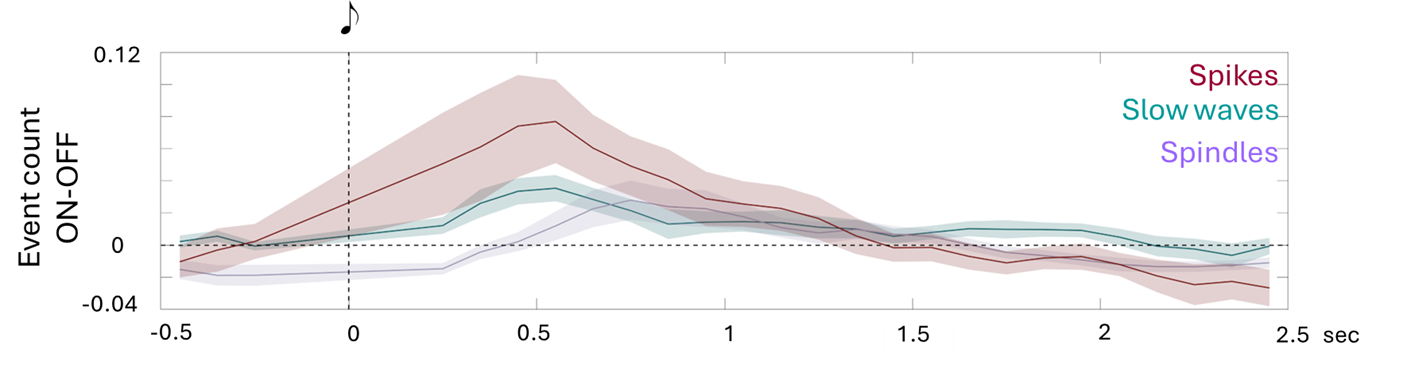

**Supplementary Figure 3 - Spikes, spindles, and slow waves over time after isolated stimuli without spike-wave complexes**

Spikes and spindles and slow waves are detected and counted like in *Figure 2*. To exclude that the slow wave response is driven by spike-wave complexes, slow waves with a negative peak occurring within 0.5 s of a detected spike were excluded (Bölsterli et al., 2011). Solid lines (red = spikes; blue = slow waves, purple = spindles) show the participant-averaged time course difference between ON and OFF stimuli. The shaded areas indicate the standard error of the mean. Spikes are shown in the predefined ROI. Spindles and slow waves are shown in Fz/Cz for isolated stimuli, as it was previously shown that this area shows the biggest stimulation effect , and in the ROI for trains of stimuli, as we expected a local effect. Outcome of the model with the differently selected slow waves can be found in *Supplementary table 15,16.*

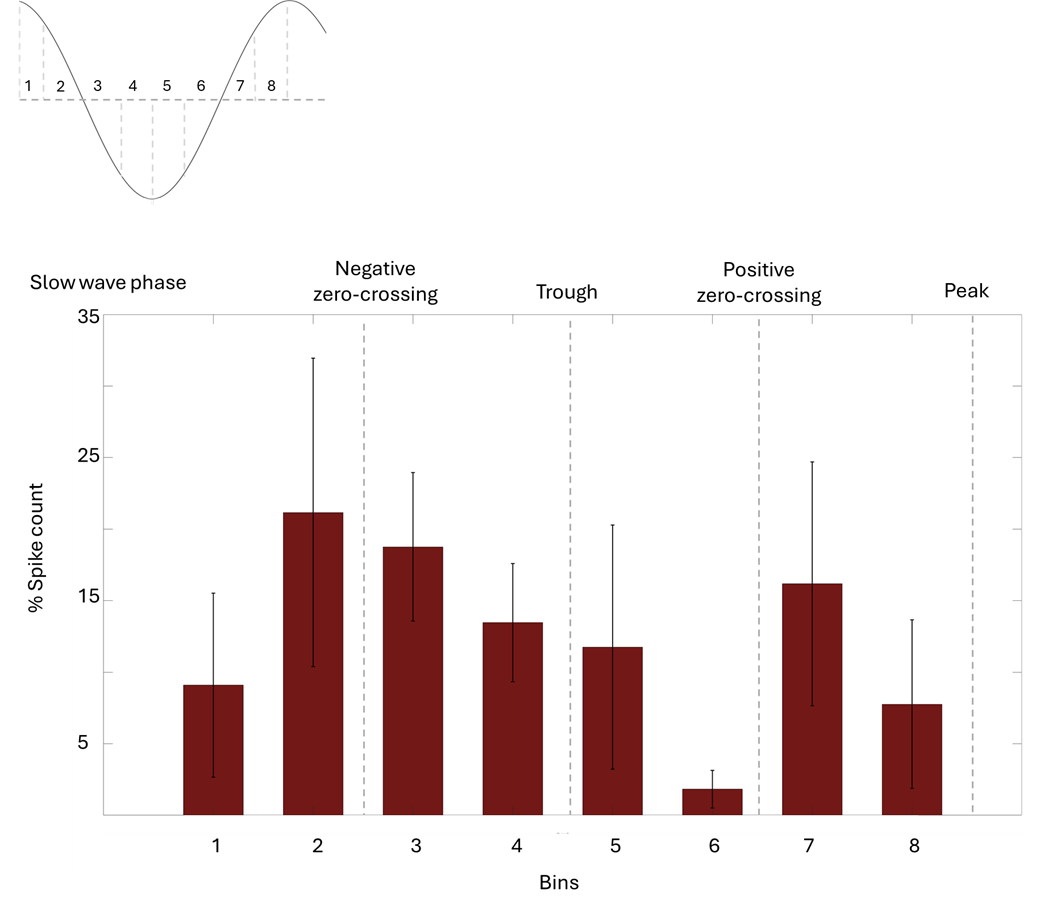

**Supplementary Figure 4 – Spikes coupling to the down-phase of evoked slow waves in cohort A**

In cohort A, big KC-like slow waves were considered to be evoked when they happened 200-800 ms after an isolated stimulus. The evoked slow waves in the ROI were then divided into eight bins between the two positive peaks, as shown in the upper left corner (see methods for more details). Spikes were assigned to bins according to their occurrence time and normalized to the duration of each bin. For each subject, spike rates were summed across evoked slow waves and normalized to the subject's total spike rate across the eight bins. Normalized spike rates were then averaged across subjects. Bars represent mean relative spike activity in the ROI ± SEM.

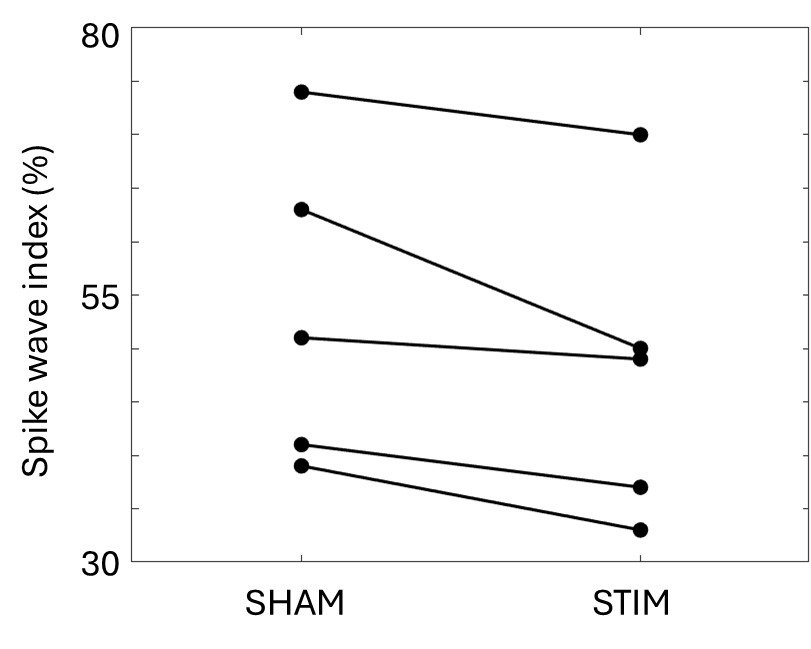

**Supplementary Figure 5 - Spike wave index comparison between stimulation and Sham night in cohort B**

A trained technician, blinded to the condition, visually detected spikes for Cohort B in a bipolar montage in 10 second epochs. This was done for 10 min of NREM sleep in the first cycle. The visual detection was done for a focus channel, chosen as the channel with the biggest automatically detected difference in spike density and closest to the detection electrode. A spike–wave index (SWI) was then calculated by dividing the number of seconds affected by spike waves by the number of seconds analysed, multiplied by 100 (Aeby et al., 2005). Each dot represents the SWI value of a patient in the Sham night, connected with a line to the respective value in the Stim night.

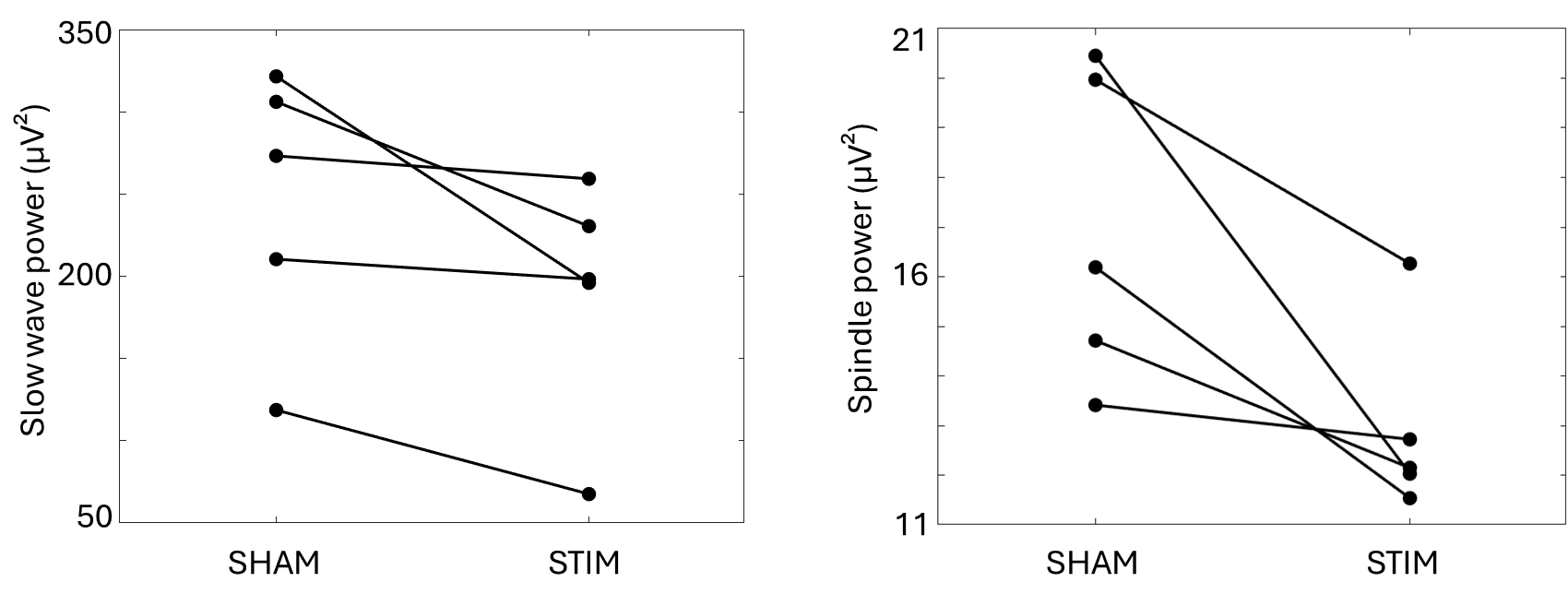

**Supplementary Figure 6 – Slow wave and spindle power comparison between stimulation and Sham night in cohort B**

Power spectral density (PSD) values were computed using Welch’s method. For slow-wave activity, power was calculated in the 0.5-4 Hz range, whereas spindle power was calculated in the 9-14 Hz range. Epoch-wise bandpower values were averaged across all included epochs, resulting in one bandpower value per channel for each participant and condition. For each patient the value in the detection channel was then extracted. Each dot represents the power value of a patient in the Sham night, connected with a line to the respective value in the Stim night (slow wave power on the left and spindle power on the right).
